# Cluster-weighted modified Poisson regression for estimating risk ratios in longitudinal data with informative cluster sizes

**DOI:** 10.1101/2025.05.23.25328253

**Authors:** Jemar R. Bather, Samuel Anyaso-Samuel, Yuyu Chen, Luther Elliott, Alex S. Bennett, Melody S. Goodman

## Abstract

Variation in binary outcomes over time by cluster size arises across various biomedical disciplines, including reproductive health, dental medicine, and psychiatric epidemiology. This study formally integrates modified Poisson regression with cluster-weighted generalized estimating equations (MP-CWGEE) for computing risk ratios in longitudinal studies with informative cluster sizes. Using a comprehensive Monte-Carlo simulation study, we empirically evaluated MP-CWGEE’s statistical properties against alternative modeling approaches: MP-GEE, log-binomial CWGEE (LB-CWGEE), and log-binomial GEE (LB-GEE). We conducted 1,000 simulations across varying sample sizes, risk ratios, and informativeness degrees. MP-CWGEE demonstrated superior performance in model convergence, empirical bias, average estimated standard error, coverage, and Type 1 error control. While LB-CWGEE showed comparable results, its convergence rates were slightly inferior. The benefits of cluster-weighted models (MP-CWGEE and LB-CWGEE) over unweighted models (MP-GEE and LB-GEE) were pronounced in scenarios with informative cluster sizes. We demonstrated MP-CWGEE’s practical application to a cohort study of people who used illicit opioids in New York City. We also provided implementation code for R, Stata, and SAS to facilitate wider adoption of the MP-CWGEE approach.

## 1 Introduction

Quantifying the association between an exposure and a binary outcome is a common goal of biomedical researchers analyzing longitudinal data [1]. While the odds ratio is commonly used to characterize this association, it has two major limitations. First, it is frequently misinterpreted as the risk ratio (or relative risk) [2], providing an accurate approximation only when the outcome prevalence is rare (<10%) [1]. Second, the odds ratio is noncollapsible (sensitive to unmodelled covariates), complicating its interpretation and comparison across studies [3, 4].

An alternative to the logit link function—which estimates the odds ratio in logistic regression— is the log link function, which directly estimates the risk ratio and provides a clinically meaningful association measure in longitudinal studies [1, 5]. Risk ratios can be calculated using log-binomial regression via maximum likelihood (ML) estimation in a mixed-effects model [6]. This is a parametric approach with a cluster-specific interpretation (conditional on the random effect) [6]. Risk ratios can also be generated using log-binomial regression via generalized estimating equations (GEE), a semi-parametric marginal modeling technique with a population-averaged interpretation [7]. However, log-binomial regression frequently encounters convergence problems [5, 8, 9]. Zou and Donner [8] addressed this limitation by developing the modified Poisson (or robust Poisson) regression model using GEE (MP-GEE). This method employs the log link function to model binary outcomes as a function of explanatory variables while using a robust variance estimator to correct the standard error overestimation that would otherwise occur in conventional Poisson regression [8].

Simulation studies have demonstrated that the MP-GEE possesses desirable statistical properties [5, 8–16]. MP-GEE exhibits greater robustness against model misspecification and outliers than the log-binomial ML model [8, 9, 11–13]. Additional simulation studies confirm that MP-GEE maintains adequate coverage, effectively controls type 1 error rates, and preserves asymptotic efficiency when estimating risk ratios [5, 9, 10]. However, despite these advantages, no published studies evaluate the performance of MP-GEE when cluster sizes are associated with the outcome (referred to as informative cluster size [17, 18]).

Outcome variation by cluster size arises across various biomedical disciplines, including sexual health [19], reproductive science [20–22], dental medicine [23, 24], and psychiatric epidemiology [25]. For example, women with multiple pregnancies face higher risks of subsequent pregnancy complications than those with a single pregnancy [26]. Analytical approaches that restrict the dataset to one pregnancy per person discard valuable information [17, 27]. Conversely, including all available data without accounting for cluster informativeness overweights individuals contributing more pregnancies, which biases regression parameter estimates [17, 27]. Two models have been developed to address this issue: within-cluster resampling and cluster-weighted generalized estimating equations (CWGEE). The within-cluster resampling approach randomly selects one observation per individual, fits a regression model, and then averages results across iterations to obtain parameter estimates [27]. While statistically valid, this method is computationally intensive. To overcome this limitation, Williamson and colleagues developed CWGEE [17], which inversely weights the GEE by cluster size. Williamson et al. [17] demonstrated that CWGEE is asymptotically equivalent to within-cluster resampling and requires less computational resources.

Combining modified Poisson regression with cluster-weighted generalized estimating equations (MP-CWGEE) is a natural extension that can account for informative cluster sizes when computing risk ratios in longitudinal studies. However, a systematic evaluation of its statistical properties has yet to be reported despite its increasing application in recent studies [28–32]. Therefore, we evaluated MP-CWGEE’s performance in estimating risk ratios in longitudinal data with informative cluster sizes.

The structure of this paper is as follows. Section 2 describes the mathematical notation, model specification, parameter interpretation, and robust variance estimator. Section 3 characterizes the simulation study comparing the MP-CWGEE model to alternative models for estimating risk ratios in longitudinal settings. Section 4 presents a real-world data application of the MP-CWGEE approach to a cohort study of people who use illicit opioids in New York City. Section 5 offers concluding remarks and discusses future research directions. Section 6 provides example codes for fitting the MP-CWGEE model in R, Stata, and SAS.

## 2 Modified Poisson regression using cluster-weighted generalized estimating equations

### 2.1 Model specification and parameter interpretation

Let *Y*_*ij*_ denote the binary outcome for the *i*^th^ individual (*i* = 1, …, *N*) on the *j*^th^ occasion (*j* = 1, …, *n*_*i*_). The total number of repeated measures for the *i*^th^ individual is reflected by *n*_*i*_ (cluster size), which can vary across study participants. Using the marginal modeling framework [7], we model the dichotomous response as a log-linear function of *p* explanatory variables:

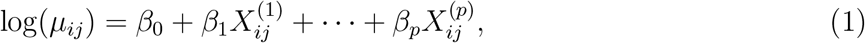

where 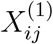 is the primary exposure of interest, 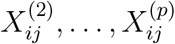 represent the covariates, and *µ*_*ij*_ = E(*Y*_*ij*_ | **X**_**ij**_) is the marginal expectation of the binary outcome. The parameter exp (*β*_1_) represents the ratio of the risk in the exposed group to that of the unexposed group, after adjusting for covariates.

### 2.2 Cluster-weighted estimation

#### 2.2.1 Parameter estimation

The MP-CWGEE estimator of the regression coefficient vector, ***β*** = (*β*_0_, *β*_1_, …, *β*_*p*_)^*T*^, is obtained by solving the following score equation [17]:

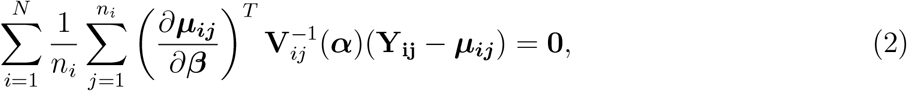

where ***α*** is the within-person correlation, **R**(***α***) is the working correlation matrix, 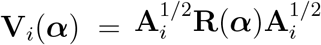 is the working variance-covariance matrix, **A**_*i*_ is the diagonal matrix with elements *ϕv*(*µ*_*ij*_), *ϕ* is the dispersion parameter, and *v*(*µ*_*ij*_) = *µ*_*ij*_ is the variance function. The inverse weighting by the cluster size (1*/n*_*i*_) ensures that each study participant contributes equally to the estimation, regardless of their number of repeated measurements [17, 19]. We assumed an independent working correlation structure, aligning with the seminal CWGEE paper [17], the seminal longitudinal modified Poisson regression paper [8], and the applied MP-CWGEE literature [31, 32]. Assuming a non-independent correlation structure (e.g., exchangeable or first-order autoregressive) requires a generalized weight: [**1**^*T*^ **R**(***α***)**1**]^*−*1^, where **1** represents an *n*_*i*_ *×* 1 vector of 1s [17, 19]. The asymptotic normality of ***β*** is derived in Section 6.2.

#### 2.2.2 Variance-covariance estimation

The robust (sandwich) variance estimator for ***β*** takes the following form:

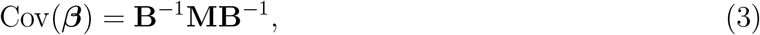

where

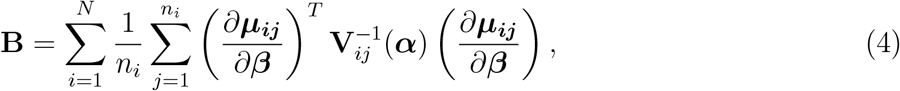

and

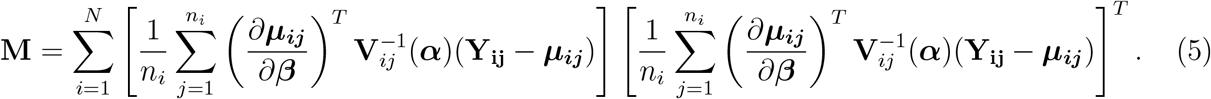

## 3 Simulation study

### 3.1 Design

We conducted a simulation study to evaluate the performance of various regression approaches for estimating risk ratios in longitudinal data with informative cluster sizes. For the *i*th subject and the *j*th observation, a binary outcome was generated with probability:

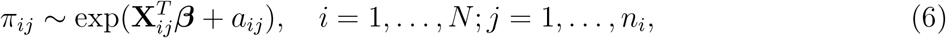

where 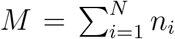 is the total number of observations. We defined the vector of explanatory variables as 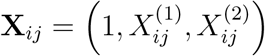, with 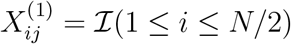 denoting a binary exposure where the first *N/*2 individuals were exposed, and 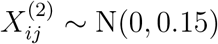 reflecting a continuous covariate. We set the true regression coefficients as ***β*** = (*β*_0_, *β*_1_, *β*_2_)^*T*^. We set *β*_0_ = log(0.37) and *β*_2_ = log(0.50).

Cluster sizes *n*_*i*_ were randomly drawn from {2, 4, 15}, with probabilities: ℙ (*n*_*i*_ = 2) = 0.5625, ℙ (*n*_*i*_ = 4) = 0.3750, and ℙ (*n*_*i*_ = 15) = 0.0625. This formulation ensures that the average number of repeated measures is approximately three. Each simulated dataset was analyzed using the following five models: MP-CWGEE, MP-GEE, log-binomial regression using cluster-weighted generalized estimating equations (LB-CWGEE), log-binomial regression using generalized estimating equations (LB-GEE), and log-binomial mixed effects model (LB-MM). Both GEE and CWGEE estimators used an independence working correlation structure.

### 3.2 Scenarios and performance metrics

We explored a range of data-generating scenarios by varying the number of individuals, the risk ratio, and the informativeness of cluster sizes. The number of individuals, *M*, was 50, 100, and 200. The risk ratios were 1.2 (modest effect) and 1.5 (moderate effect). For the informativeness of cluster sizes, we defined

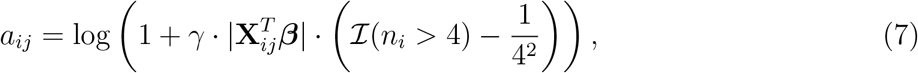

where *γ* ∈ {0, 1, 1.5} controls the degree of informativeness [33]. Note that *γ* = 0 indicates non-informative cluster sizes. Using 1,000 Monte-Carlo simulations, we evaluated the following performance metrics: model convergence, empirical bias, average estimated standard error, coverage rates, and empirical Type 1 error rates.

### 3.3 Simulation study results

#### 3.3.1 Model convergence

Table 1 shows the convergence rates of all five models across all simulation settings. MP-CWGEE and MP-GEE consistently achieved perfect convergence (100%) across all simulation scenarios, regardless of the degree of informativeness, exposure effect, and sample size. LB-CWGEE also performed well, with convergence rates of at least 99%. LB-GEE exhibited reasonable convergence rates (all >88%), but these rates decreased as sample size decreased and informativeness increased. LB-MM demonstrated poor convergence across all settings (<36%), with the lowest rate (<1%) in a scenario with 200 study participants, a 1.5 exposure effect, and informativeness = 1.5. Given LB-MM’s poor convergence, we do not discuss its other performance metrics (e.g., bias) in the remainder of this report as these metrics would not be comparable to those of the models that showed good convergence. The non-convergence of the LB-MM was likely due to maximum likelihood estimates being on the parameter space boundary [11, 34].

**Table 1.** Model convergence rates (%) by sample size, exposure effect, and informativeness degree.

| N | Risk ratio | Informativeness ($\gamma$) | MP-CWGEE | MP-GEE | LB-CWGEE | LB-GEE | LB-MM |
| --- | --- | --- | --- | --- | --- | --- | --- |
| 50 | 1.2 | 0.0 | 100.0 | 100.0 | 99.3 | 96.1 | 34.2 |
|  |  | 1.0 | 100.0 | 100.0 | 99.6 | 94.1 | 35.1 |
|  |  | 1.5 | 100.0 | 100.0 | 99.8 | 92.9 | 22.3 |
|  | 1.5 | 0.0 | 100.0 | 100.0 | 99.1 | 95.4 | 19.4 |
|  |  | 1.0 | 100.0 | 100.0 | 99.4 | 93.3 | 17.5 |
|  |  | 1.5 | 100.0 | 100.0 | 99.0 | 88.9 | 10.7 |
| 100 | 1.2 | 0.0 | 100.0 | 100.0 | 100.0 | 99.3 | 24.9 |
|  |  | 1.0 | 100.0 | 100.0 | 100.0 | 98.4 | 25.0 |
|  |  | 1.5 | 100.0 | 100.0 | 100.0 | 97.8 | 14.1 |
|  | 1.5 | 0.0 | 100.0 | 100.0 | 99.9 | 98.1 | 6.6 |
|  |  | 1.0 | 100.0 | 100.0 | 99.6 | 96.6 | 5.8 |
|  |  | 1.5 | 100.0 | 100.0 | 99.7 | 95.7 | 2.8 |
| 200 | 1.2 | 0.0 | 100.0 | 100.0 | 100.0 | 99.6 | 16.7 |
|  |  | 1.0 | 100.0 | 100.0 | 100.0 | 99.2 | 16.2 |
|  |  | 1.5 | 100.0 | 100.0 | 100.0 | 99.7 | 7.2 |
|  | 1.5 | 0.0 | 100.0 | 100.0 | 99.9 | 99.0 | 1.9 |
|  |  | 1.0 | 100.0 | 100.0 | 100.0 | 98.6 | 1.7 |
|  |  | 1.5 | 100.0 | 100.0 | 100.0 | 98.0 | 0.9 |

MP-CWGEE = modified Poisson regression using cluster-weighted generalized estimating equations
MP-GEE = modified Poisson regression using (unweighted) generalized estimating equations
LB-CWGEE = log-binomial regression using cluster-weighted generalized estimating equations
LB-GEE = log-binomial regression using (unweighted) generalized estimating equations
LB-MM = log-binomial mixed effects model

#### 3.3.2 Empirical bias

Fig. 1 illustrates the empirical bias of *β*_1_ across varying informativeness degrees and sample sizes. When cluster sizes were non-informative (*γ* = 0), all four models showed comparable empirical bias patterns, regardless of sample size or *β*_1_ value. However, with informative cluster sizes (*γ >* 0), the CWGEE approaches (MP-CWGEE and LB-CWGEE) produced substantially less biased estimates than their unweighted counterparts.

**Figure 1.**
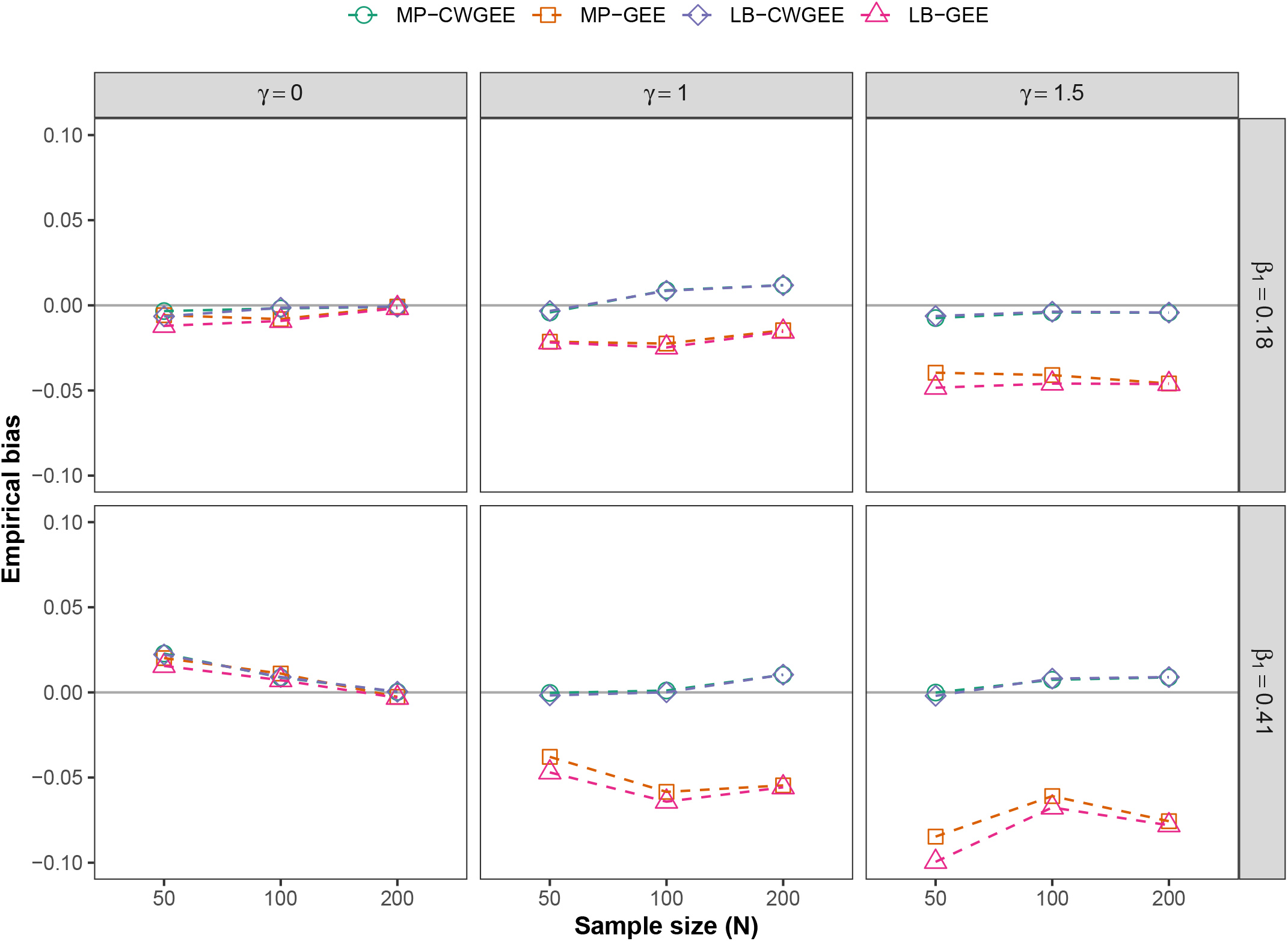
Comparison of bias in parameter estimates for four risk ratio estimation models under varying conditions. The *γ* parameter reflects the informativeness degree (*γ* = 0 is non-informative). Note that exp(0.18) = 1.2 and exp(0.41) = 1.5, denoting the true risk ratios. MP-CWGEE = modified Poisson regression using cluster-weighted generalized estimating equations; MP-GEE = modified Poisson regression using (unweighted) generalized estimating equations; LB-CWGEE = log-binomial regression using cluster-weighted generalized estimating equations; LB-GEE = log-binomial regression using (unweighted) generalized estimating equations.

#### 3.3.3 Average estimated standard error

Fig. 2 visualizes patterns in the average estimated standard error across 1,000 Monte-Carlo simulations. For all four models, regardless of informativeness degree and magnitude of *β*_1_, standard errors decreased as the sample size increased. At *γ* = 0 (non-informative cluster sizes), unweighted models (MP-GEE and LB-GEE) had smaller standard errors than cluster-weighted models (MP-CWGEE and LB-CWGEE). This trend eventually reversed in the presence of informative cluster sizes (*γ* > 0), with cluster-weighted models showing better precision than unweighted models.

**Figure 2.**
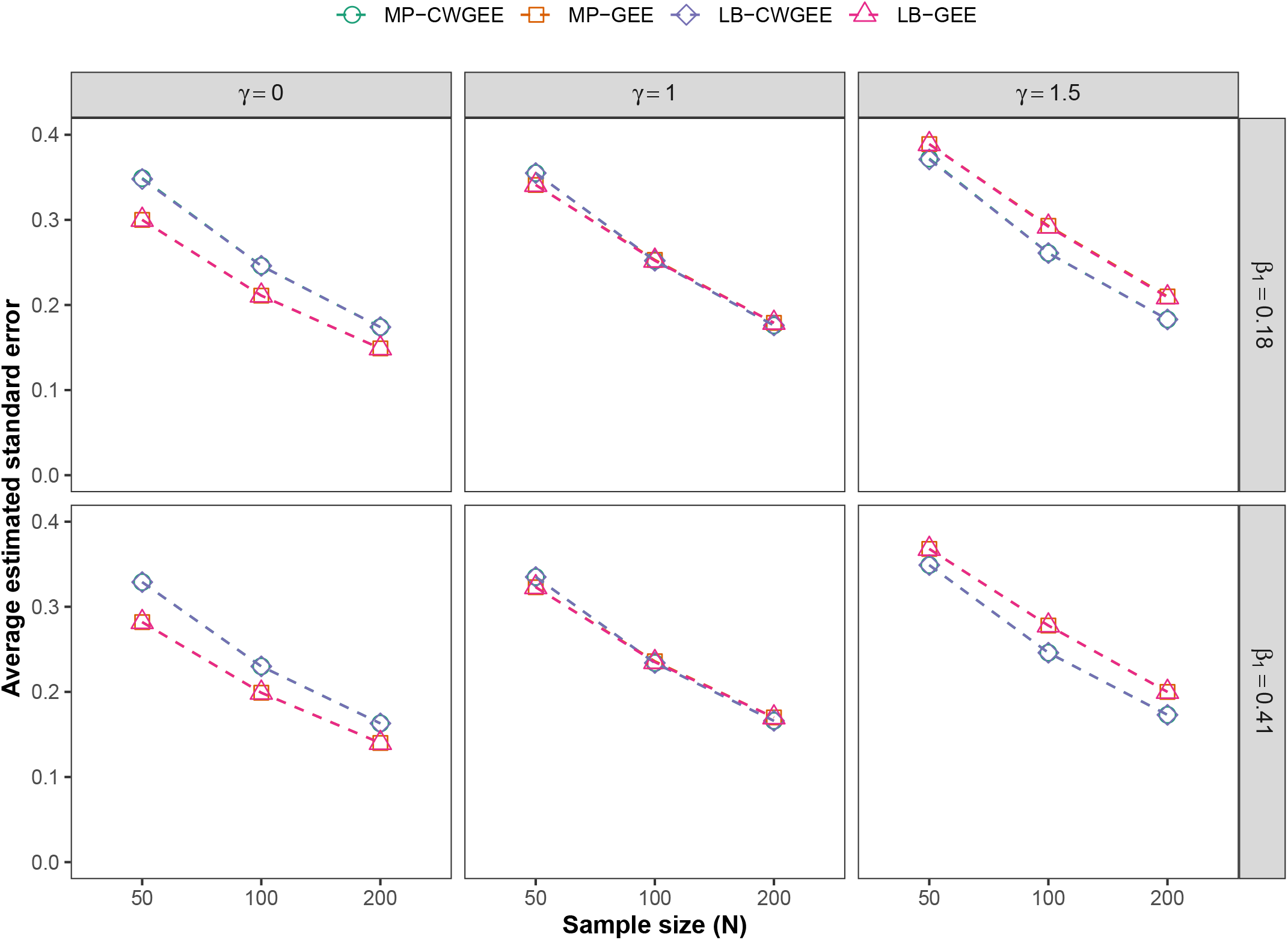
Average estimated standard error comparison across four risk ratio estimation models under varying conditions. The *γ* parameter reflects the informativeness degree (*γ* = 0 is non-informative). Note that exp(0.18) = 1.2 and exp(0.41) = 1.5, denoting the true risk ratios. MP-CWGEE = modified Poisson regression using cluster-weighted generalized estimating equations; MP-GEE = modified Poisson regression using (unweighted) generalized estimating equations; LB-CWGEE = log-binomial regression using cluster-weighted generalized estimating equations; LB-GEE = log-binomial regression using (unweighted) generalized estimating equations.

#### 3.3.4 Coverage

Table 2 summarizes the coverage rates for *β*_1_ by sample size, exposure effects, and informativeness degree. Coverage rates decreased for the unweighted models as the informativeness parameter, *γ*, increased from 0.0 to 1.5. For example, at N = 50 and risk ratio = 1.2, the MP-GEE coverage decreased from 93.9% (*γ* = 0.0) to 88.9% (*γ* = 1.0) to 86.9% (*γ* = 1.5). Increasing the sample size mitigated these coverage issues. For instance, at *γ* = 1.5 and risk ratio = 1.2, MP-GEE coverage improved from 86.9% (N = 50) to 91.0% (N = 100) to 93.1% (N = 200). By contrast, the cluster-weighted approaches maintained coverage rates close to the nominal 95% level across all simulated scenarios.

**Table 2.**
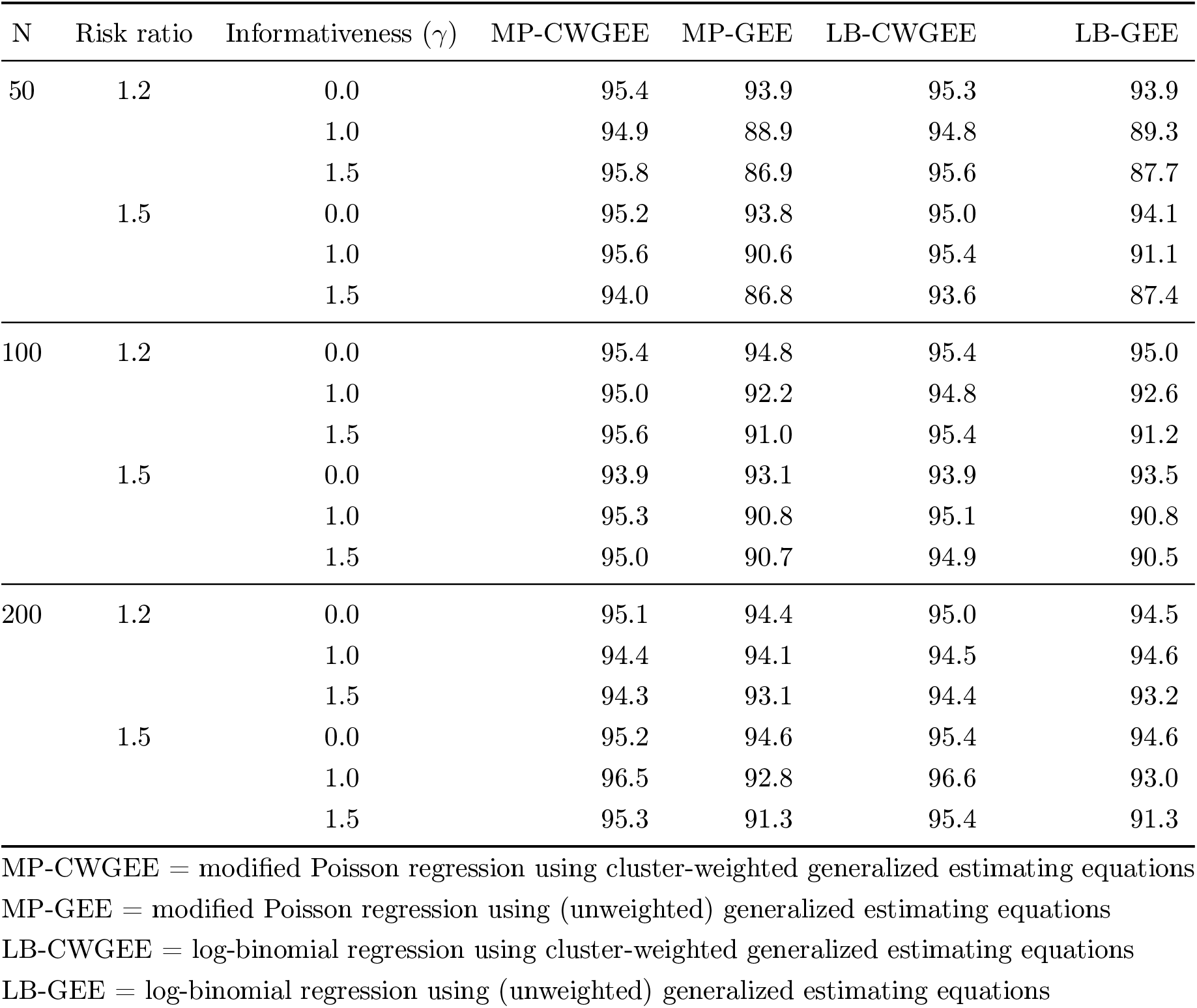
Coverage rates (%) for *β*_1_ across different simulation settings.

#### 3.3.5 Empirical Type 1 error rate

Fig. 3 summarizes the results from the analysis of Type 1 error rates. When *γ* = 0 (uninformative cluster sizes) and N = 50, all models yielded slightly inflated Type 1 error rates (range: 6.3-7.0%), but this improved as sample size increased to 100 (range: 3.6-4.7%) and 200 (range: 3.9-4.8%). As informativeness increased, the unweighted models exhibited inflated Type 1 error rates, particularly at smaller sample sizes. For example, at *γ* = 1.5 and N = 50, MP-GEE had a 12.5% Type 1 error rate, and LB-GEE had an 11.9% Type 1 error rate. In contrast, the cluster-weighted models controlled the Type 1 error rate as informativeness increased.

**Figure 3.**
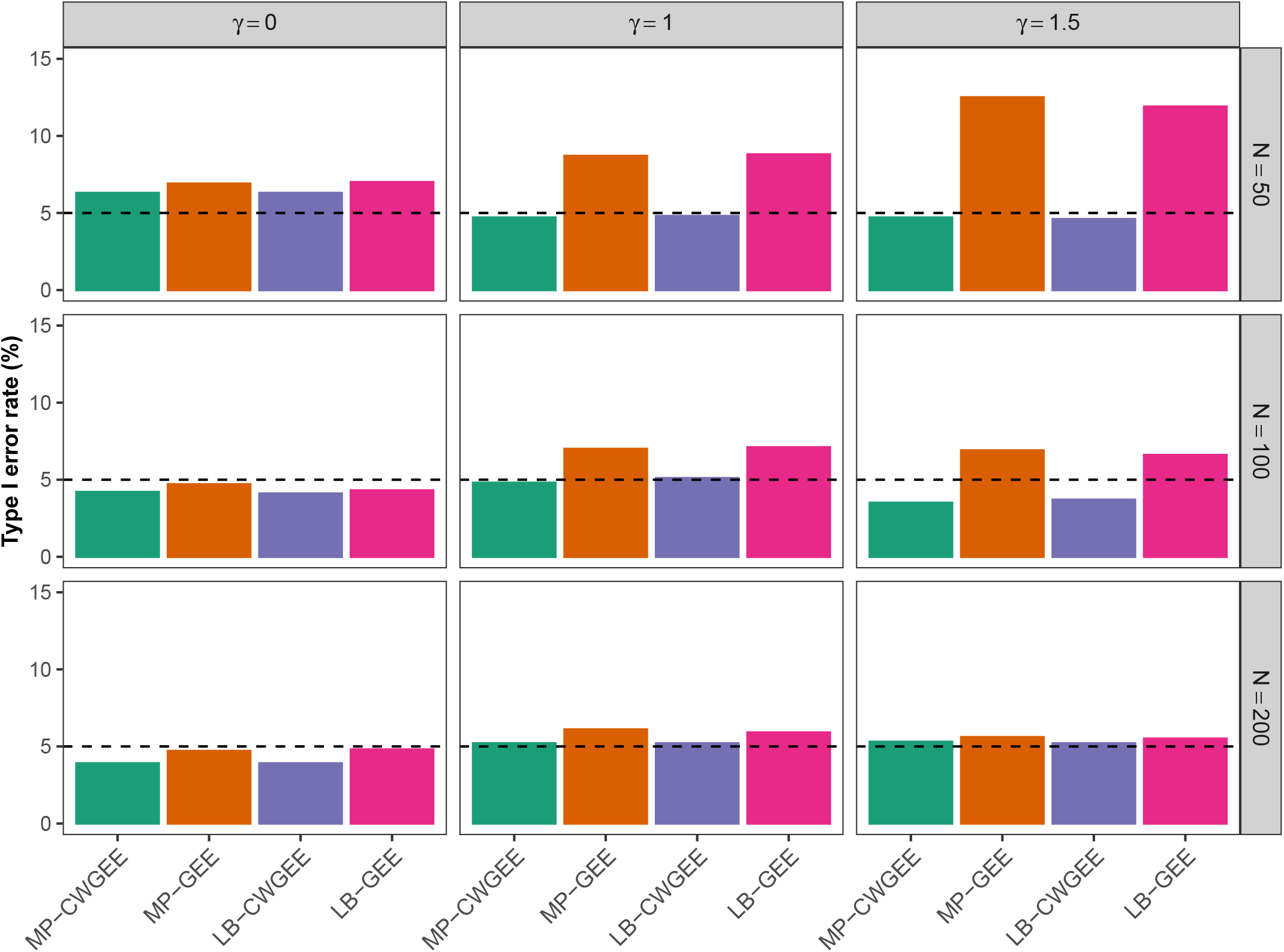
Empirical Type 1 error rates for four risk ratio estimation models under varying conditions. The *γ* parameter reflects the informativeness degree (*γ* = 0 is non-informative). MP-CWGEE = modified Poisson regression using cluster-weighted generalized estimating equations; MP-GEE = modified Poisson regression using (unweighted) generalized estimating equations; LB-CWGEE = log-binomial regression using cluster-weighted generalized estimating equations; LB-GEE = log-binomial regression using (unweighted) generalized estimating equations.

#### 3.3.6 Additional simulation results

The simulation results presented up to this point primarily focus on the performance of the estimators for *β*_1_, the exposure effect. However, prior studies [23, 35] have shown that, when informative cluster sizes are not properly accounted for, the estimation of other regression parameters can also be biased. To provide a more comprehensive evaluation of the regression models, we report simulation results for the estimation of the intercept *β*_0_ and the continuous covariate effect *β*_2_.

Supplemental Figures 1 and 2 in the Supplementary Material present the relative bias of the estimators for *β*_0_ and *β*_2_, respectively, across varying levels of informativeness and sample sizes. For both regression parameters, the results reveal consistent patterns across simulation settings. When cluster sizes are non-informative, all models yield approximately unbiased estimates, with bias decreasing as the number of samples increases. In contrast, under informative cluster size scenarios, the GEE models exhibit substantial bias, particularly for *β*_0_. The CWGEE models are markedly more robust to informativeness, with bias decreasing as sample size grows. Conversely, the bias in the GEE estimators tends to worsen as the number of samples increases.

Supplemental Figures 3 and 4 of the Supplementary Material show the average estimated standard errors for *β*_0_ and *β*_2_, respectively. Across all scenarios, GEE-based models report smaller standard errors than their CWGEE counterparts, irrespective of the degree of informativeness. This reflects the well-known trade-off between bias and variance in model estimation [36].

Coverage probabilities for *β*_0_ and *β*_2_ are reported in Supplemental Tables 1 and 2. For *β*_2_, all models achieve coverage rates close to the nominal 95% level, even under informative cluster sizes. However, for *β*_0_, the GEE models consistently fall short of the nominal coverage level when cluster sizes are informative. Notably, this undercoverage does not improve with increasing sample size. In contrast, the CWGEE models maintain coverage rates near 95%, further underscoring their robustness in the presence of informativeness.

## 4 Application to a study of people who use illicit opioids in New York City

### 4.1 Data source

We analyzed repeated measures data from a prospective cohort study of over 400 adults (aged 18+) who used illicit opioids in New York City [37, 38]. The dataset comprised 7,365 observations on 423 adults collected during monthly study engagements between 2019 and 2022. Complete details regarding the study design and recruitment procedures are described elsewhere [37, 38]. The New York University Grossman School of Medicine Institutional Review Board approved this study protocol.

### 4.2 Measures

The binary outcome of interest was whether the participant had any clinical indicators of major depressive disorder (1 = any, 0 = none). This was measured by the Cross-Cutting Depression Sub-scale from the Diagnostic and Statistical Manual of Mental Disorders Version 5 [39]. Race/ethnicity was the independent variable of interest and categorized as non-Hispanic White, non-Hispanic Black, Hispanic, and non-Hispanic Other. Covariates included age (measured continuously), educational attainment (did not complete high school, high school diploma/GED, some college or more), and having an opioid-using network size *≥*5 (yes vs. no).

### 4.3 Statistical analysis

Exploratory data analysis indicated that study participants had variable engagement with the research protocol, with considerable heterogeneity in study engagements (mean: 17 engagements, SD: 7, Table 3). Given the well-established clinical observation that individuals experiencing depression are less likely to participate in follow-up protocols [40, 41], we hypothesized that the number of study engagements (cluster size) would be informative of the outcome. To test this hypothesis, we modeled having clinical indicators of major depressive disorder as a function of study engagement frequency (3-4 engagements, 5-10 engagements, 11-16 engagements, 17-24 engagements) using an MP-GEE model with an independent correlation structure. Cut-offs for the engagement frequency variable were derived based on exploratory analyses and subject matter expertise.

**Table 3.** Baseline characteristics of 423 adults who used illicit opioids in New York City.

| Characteristic | N = 423 |
| --- | --- |
| Study engagements, Mean (SD) | 17 (7) |
| Age, Mean (SD) | 48 (11) |
| Has clinical indicators of major depressive disorder, No. (%) |  |
| No | 194 (46%) |
| Yes | 229 (54%) |
| Race/ethnicity, No. (%) |  |
| non-Hispanic White | 82 (19%) |
| non-Hispanic Black | 148 (35%) |
| Hispanic | 173 (41%) |
| non-Hispanic Other | 20 (5%) |
| Educational attainment, No. (%) |  |
| Did Not Complete High School | 102 (24%) |
| High School or GED Graduate | 173 (41%) |
| Some College or More | 148 (35%) |
| Has an opioid-using network size $\geq 5$, No. (%) | |
| No | 101 (24%) |
| Yes | 322 (76%) |

To quantify the adjusted association between race/ethnicity and having clinical indicators of major depressive disorder, we compared adjusted risk ratios (and 95% CIs) across different statistical models: MP-CWGEE, MP-GEE, LB-CWGEE, LB-GEE, and LB-MM. The MP-CWGEE and LB-CWGEE models were weighted by the inverse of study engagements (measured continuously). All models controlled for age, educational attainment, and having an opioid-using network size ≥5. Analyses used R version 4.4.3 (R Core Team, R Foundation for Statistical Computing), with statistical significance assessed as a 2-sided *p*<0.05.

### 4.4 Results

Table 3 displays the baseline characteristics of the study population. Of the 423 participants (mean age: 48 years, SD: 11), 54% had clinical indicators of major depressive disorder, 41% identified as Hispanic, 24% did not graduate high school, and 76% had an opioid-using network size ≥5. Table 4 presents the associations between study engagement frequency and having clinical indicators of major depressive disorder. Compared to those with 3-4 study engagements, individuals with 11-16 study engagements were 30% (RR: 0.70, 95% CI: 0.51-0.95, *p*=0.024) less likely to have clinical indicators of major depressive disorder, indicating nonignorable cluster size. Individuals with 17-24 study engagements were 21% (RR: 0.79, 95% CI: 0.62-1.02, *p*=0.068) less likely to have clinical indicators of major depressive disorder than those with 3-4 study engagements, but this association was not statistically significant.

**Table 4.** Associations between study engagement frequency and having clinical indicators of major depressive disorder among 423 adults who used illicit opioids in New York City.

| Characteristic | RR | 95% CI | p-value |
| --- | --- | --- | --- |
| Number of study engagements |  |  |  |
| 3-4 engagements | — | — |  |
| 5-10 engagements | 0.78 | 0.55, 1.11 | 0.20 |
| 11-16 engagements | 0.70 | 0.51, 0.95 | 0.024 |
| 17-24 engagements | 0.79 | 0.62, 1.02 | 0.068 |

RR = Risk Ratio, CI = Confidence Interval

Table 5 presents the comparison of statistical models that quantify the adjusted associations between race/ethnicity and having clinical indicators of major depressive disorder. Consistently across all models, individuals who identified as non-Hispanic Black were significantly less likely to have clinical indicators of major depressive disorder compared to their non-Hispanic White counterparts: MP-CWGEE (RR: 0.66, *p*=0.004), MP-GEE (RR: 0.67, *p*=0.012), LB-CWGEE (RR: 0.67, *p*=0.005), LB-GEE (RR: 0.67, *p*=0.013). The cluster-weighted models yielded narrower 95% CIs, reflecting greater precision in their estimates relative to unweighted models: MP-CWGEE (95% CI: 0.50-0.88) vs. MP-GEE (95% CI: 0.49-0.92); LB-CWGEE (95% CI: 0.50-0.88) vs. LB-GEE (95% CI: 0.49-0.92). The MP-CWGEE and LB-CWGEE models produced similar risk ratios and 95% CIs, which was expected since both theoretically estimate the same association parameter. We observed a similar finding when comparing the MP-GEE and LB-GEE models’ risk ratios and 95% CIs. The LB-MM model failed to produce standard errors even after using the other models’ estimates as initial starting values.

**Table 5.**
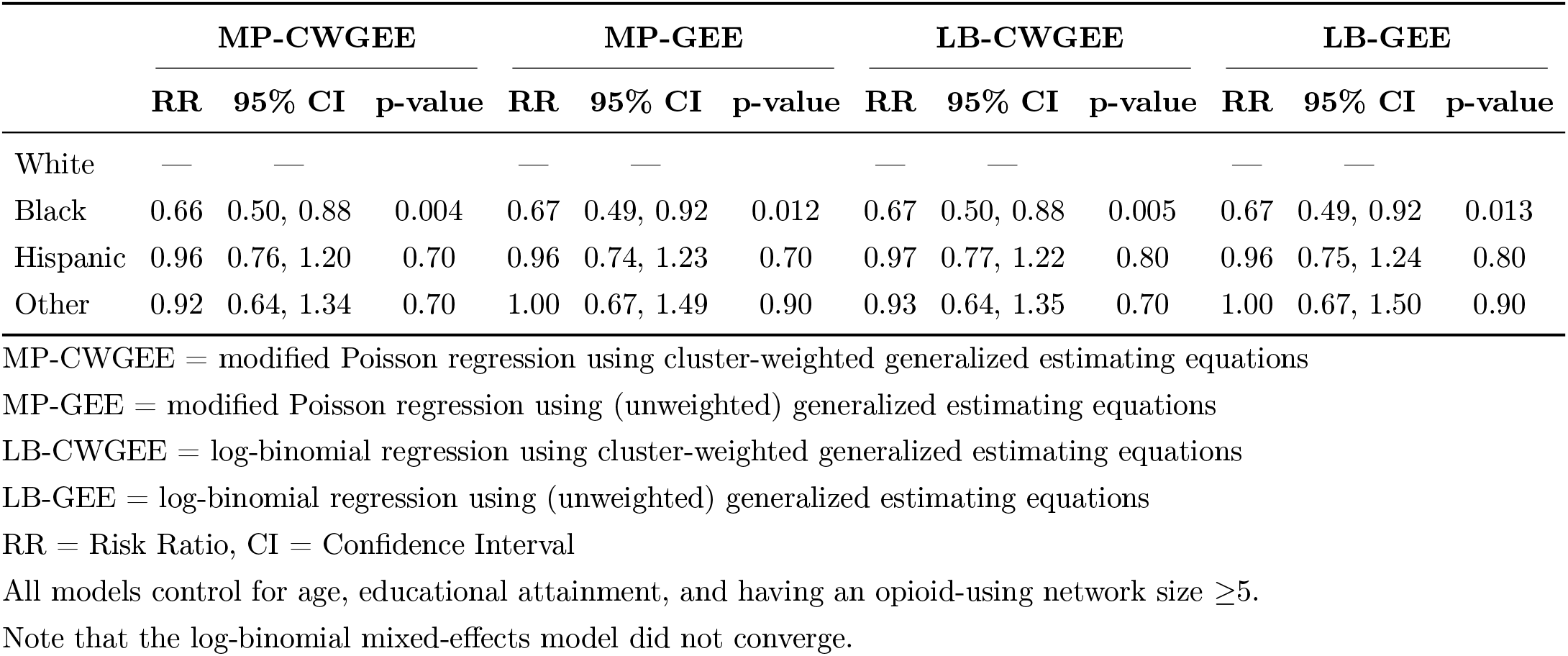
Comparison of different statistical models quantifying the adjusted associations between race/ethnicity and having clinical indicators of major depressive disorder among 423 adults who used illicit opioids in New York City.

|  | MP-CWGEE |  |  | MP-GEE |  |  | LB-CWGEE |  |  | LB-GEE |  |  |
| --- | --- | --- | --- | --- | --- | --- | --- | --- | --- | --- | --- | --- |
|  | RR | 95% CI | p-value | RR | 95% CI | p-value | RR | 95% CI | p-value | RR | 95% CI | p-value |
| White | — | — |  | — | — |  | — | — |  | — | — |  |
| Black | 0.66 | 0.50, 0.88 | 0.004 | 0.67 | 0.49, 0.92 | 0.012 | 0.67 | 0.50, 0.88 | 0.005 | 0.67 | 0.49, 0.92 | 0.013 |
| Hispanic | 0.96 | 0.76, 1.20 | 0.70 | 0.96 | 0.74, 1.23 | 0.70 | 0.97 | 0.77, 1.22 | 0.80 | 0.96 | 0.75, 1.24 | 0.80 |
| Other | 0.92 | 0.64, 1.34 | 0.70 | 1.00 | 0.67, 1.49 | 0.90 | 0.93 | 0.64, 1.35 | 0.70 | 1.00 | 0.67, 1.50 | 0.90 |

MP-CWGEE = modified Poisson regression using cluster-weighted generalized estimating equations
MP-GEE = modified Poisson regression using (unweighted) generalized estimating equations
LB-CWGEE = log-binomial regression using cluster-weighted generalized estimating equations
LB-GEE = log-binomial regression using (unweighted) generalized estimating equations
RR = Risk Ratio, CI = Confidence Interval
All models control for age, educational attainment, and having an opioid-using network size $\geq 5$ .
Note that the log-binomial mixed-effects model did not converge.

## 5 Discussion

Accounting for informative cluster sizes when computing risk ratios in longitudinal data arises in various medical research areas, including reproductive epidemiology [28, 30, 31] and infection prevention [29]. Despite its potential utility, MP-CWGEE remains underused in published studies, with limited information regarding its operating characteristics. Using a comprehensive Monte-Carlo simulation study, we empirically evaluated MP-CWGEE’s statistical properties against alternative models, including MP-GEE, LB-CWGEE, and LB-GEE. Results indicated that MP-CWGEE consistently exhibited superior performance across key metrics: model convergence, empirical bias, average estimated standard error, coverage, and Type 1 error control. While LB-CWGEE performed similarly to MP-CWGEE, it demonstrated slightly poorer convergence rates. The advantages of cluster-weighted models (MP-CWGEE and LB-CWGEE) over unweighted models (MP-GEE and LB-GEE) became particularly evident when informative cluster sizes were present.

Given that MP-CWGEE is nested within the GEE framework [7], it possesses several notable advantages over ML models. Compared to ML approaches, MP-CWGEE is less computationally intensive and requires fewer assumptions about the exposure-outcome relationship [7]. MP-CWGEE does not assume a random effect distribution, proceeds without requiring residuals to be normally distributed, and eliminates the need for correctly specifying the working correlation matrix [7]. From a health science perspective, MP-CWGEE provides additional benefits. It addresses the noncollapsibility of the odds ratio, resulting in a more clinically intuitive and meaningful association measure [1, 3, 42]. In addition, the MP-CWGEE model can be implemented in standard statistical software packages (e.g., R, SAS, Stata), making it accessible for applications across diverse epidemiologic fields such as reproductive medicine, psychiatric epidemiology, and hospital infection prevention. Sample code for fitting the MP-CWGEE model in R, SAS, and Stata is provided in Section 6.1.

Despite these strengths, MP-CWGEE has two major limitations. First, MP-CWGEE may compute predicted probabilities exceeding one [42], as the Poisson distribution’s support is non-negative. Consequently, MP-CWGEE may be more suitable for association and causal studies than prediction studies. Second, since MP-CWGEE is a semi-parametric approach, model selection cannot rely on parametric test statistics such as the likelihood ratio test, Akaike information criterion, or Bayesian information criterion [43]. In practice, subject matter expertise should guide the final model selection.

The present study lays the groundwork for future research. One priority area is empirically estimating the impact of missing data patterns (e.g., missing at random, missing not at random) and evaluating corrective approaches such as inverse probability weighting and multiple imputation within the MP-CWGEE framework. Additionally, studies are needed to assess MP-CWGEE’s sensitivity to outliers and model misspecification [11, 12]. It is unknown how robust the MP-CWGEE approach is to omitted-variable bias [44]. In other words, how does MP-CWGEE perform when an important interaction or quadratic term is not included in the model [11]? Small sample performance represents another crucial research gap. Future work should investigate finite sample bias-correction methods for MP-CWGEE when analyzing fewer than 30 clusters [45–47]. Lastly, the present study assumed that only the outcome depended on cluster size, but this relationship may be more complex. Comprehensive simulation studies should explore various potential relationships between outcomes, exposures, confounders, and cluster size.

In settings where cluster sizes are non-informative, both CWGEE and GEE models yield approximately unbiased estimates; however, GEE tends to be more efficient due to its smaller standard errors. This highlights the importance of assessing informativeness prior to model selection, as unnecessary adjustments for informativeness can lead to a loss of efficiency. Several formal statistical procedures have been developed to test for informative cluster size across a variety of outcome types, including both continuous and non-continuous data [48–52]. Future work could explore the performance and applicability of these tests specifically within the context of modified Poisson regression, where such evaluations remain limited.

## Conclusions

This study evaluated the performance of the MP-CWGEE model for estimating risk ratios in longitudinal data with informative cluster sizes. Monte-Carlo simulations demonstrated that MP-CWGEE outperformed alternative modeling approaches regarding model convergence, empirical bias, precision, coverage, and Type 1 error control. The applied example using data from adults who use illicit opioids in New York City confirmed MP-CWGEE’s utility in social epidemiologic research, providing more precise estimates than unweighted models in the presence of informative cluster sizes. Despite limitations such as potentially predicted probabilities exceeding one, MP-CWGEE offers a practical and accessible solution for analyzing longitudinal binary data with informative cluster sizes.

## Acknowledgments

We thank the study participants for contributing to the cohort study used for the data application.

## Declarations

### Author contributions

**JRB**: Conceptualization, Methodology, Software, Formal analysis, Writing (original draft), Visualization. **SA:** Conceptualization, Methodology, Software, Formal analysis, Writing (original draft), Visualization. **YC:** Investigation, Writing (review & editing). **LE:** Investigation, Writing (review & editing), Funding acquisition. **ASB:** Investigation, Writing (review & editing), Funding acquisition. **MSG:** Conceptualization, Investigation, Writing (review & editing).

### Data availability statement

The data used for the data application is available upon reasonable request.

### Ethics approval and consent to participate

The study protocol was approved by the New York University Langone Health Institutional Review Board. All study participants provided informed consent.

## 6 Appendix

### 6.1 Implementation code for R, Stata, and SAS

This section provides example code for fitting the MP-CWGEE model with an independent correlation structure using the `geepack` package in `R`, the `PROC GENMOD` procedure in `SAS`, and the `xtgee` command in `Stata`. The inverse cluster weight (one divided by the number of repeated measures) is denoted by inverse_n_measures.

#### R code

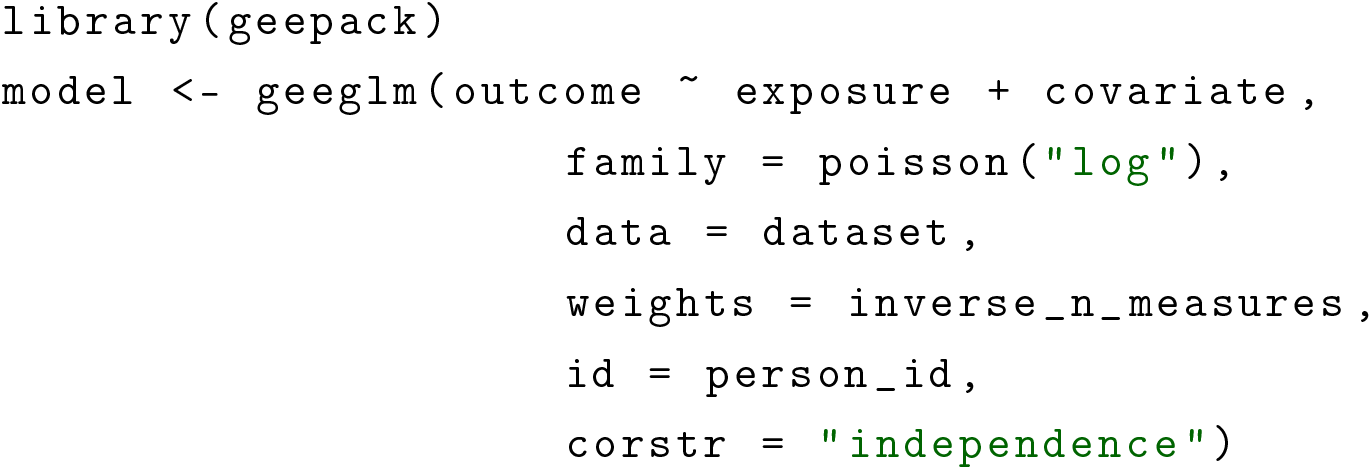

#### SAS code

~~~
PROC GENMOD DATA = dataset;
  CLASS person _id;
  MODEL outcome = exposure covariate / DIST = POISSON LINK = LOG;
  WEIGHT inverse _n_measures;
  REPEATED SUBJECT = person _id / TYPE = IND;
RUN;
~~~

#### Stata code

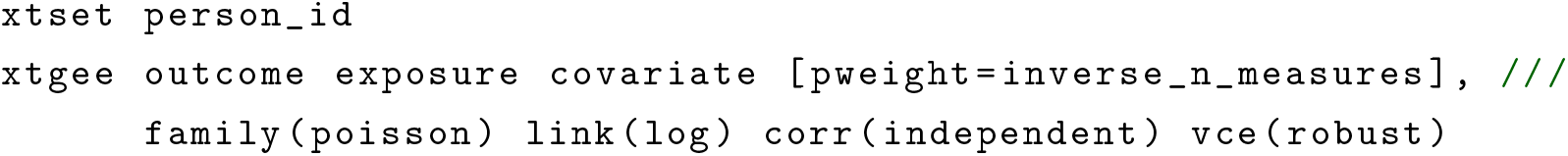

### 6.2 Asymptotic normality of MP-CWGEE estimator

#### Theorem 1.

*Under suitable regularity conditions, the MP-CWGEE estimator* 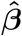 *is asymptotically normal:*

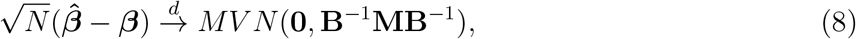

*where* ***β*** *is the vector of true parameter values*, **B** *is the limit of the expected derivative matrix, and* **M** *is the limit of the variance-covariance matrix of the estimating functions*.

*Proof*. The following proof employs a strategy and uses notation similar to Williamson et al. [17]. We consider the asymptotic distribution of the MP-CWGEE estimator 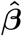 that solves the estimating equation:

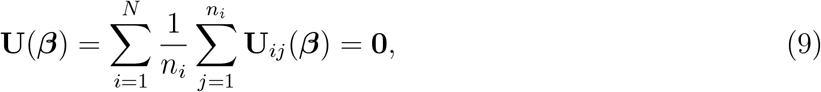

where **U**_*ij*_(***β***) represents the contribution to the score function from the *j*-th observation in the *i*-th cluster, and *n*_*i*_ is the size of cluster *i*. Define 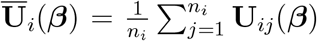 as the average score contribution from cluster *i*. Under the true parameter vector ***β***, we have 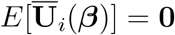. for all *i*. Let 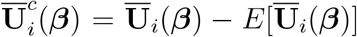 denote the centered version. Note that 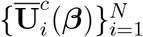 are independent with mean zero. To establish asymptotic normality, we verify the conditions for Lyapunov’s central limit theorem. For some *α >* 0, the (2 + *α*)-th moments are bounded:

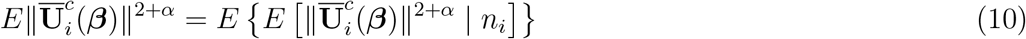

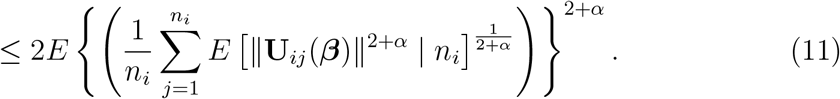

We assume that *E*[∥**U**_*ij*_(***β***)∥^2+*α*^ | *n*_*i*_] ≤ *A*_*i*_ with sup*i E*[*A*_*i*_] < *∞*, which ensures that the above expectation is bounded. We assume that the average variance-covariance matrix converges:

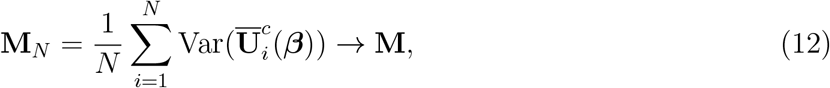

where **M** is a positive definite matrix. Note that

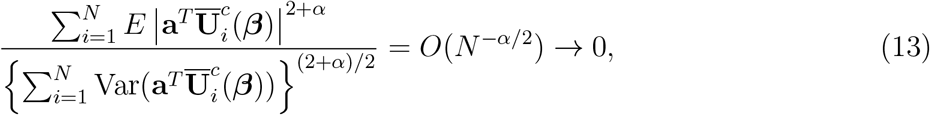

where **a** is any *d*_1_ *×* 1 vector. Consequently, we can invoke Lyapounov’s central limit theorem:

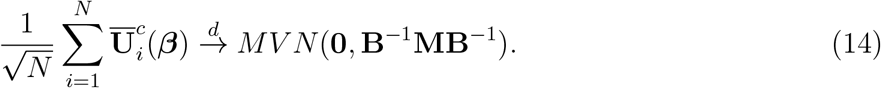

## References

1. Greenland, S. Interpretation and choice of effect measures in epidemiologic analyses. American Journal of Epidemiology 125, 761–768 (1987).

2. Holmberg, M. J. & Andersen, L. W. Estimating risk ratios and risk differences: alternatives to odds ratios. JAMA 324, 1098–1099 (2020).

3. Greenland, S., Pearl, J. & Robins, J. M. Confounding and collapsibility in causal inference. Statistical Science 14, 29–46 (1999).

4. Richardson, T. S., Robins, J. M. & Wang, L. On modeling and estimation for the relative risk and risk difference. Journal of the American Statistical Association 112, 1121–1130 (2017).

5. Zou, G. A modified poisson regression approach to prospective studies with binary data. American Journal of Epidemiology 159, 702–706 (2004).

6. Fitzmaurice, G. M., Laird, N. M. & Ware, J. H. Applied longitudinal analysis (John Wiley & Sons, 2012).

7. Liang, K.-Y. & Zeger, S. L. Longitudinal data analysis using generalized linear models. Biometrika 73, 13–22 (1986).

8. Zou, G. & Donner, A. Extension of the modified Poisson regression model to prospective studies with correlated binary data. Statistical Methods in Medical Research 22, 661–670 (2013).

9. Yelland, L. N., Salter, A. B. & Ryan, P. Performance of the modified Poisson regression approach for estimating relative risks from clustered prospective data. American Journal of Epidemiology 174, 984–992 (2011).

10. Li, F. & Tong, G. Sample size estimation for modified Poisson analysis of cluster randomized trials with a binary outcome. Statistical Methods in Medical Research 30, 1288–1305 (2021).

11. Chen, W., Qian, L., Shi, J. & Franklin, M. Comparing performance between log-binomial and robust Poisson regression models for estimating risk ratios under model misspecification. BMC Medical Research Methodology 18, 1–12 (2018).

12. Chen, W., Shi, J., Qian, L. & Azen, S. P. Comparison of robustness to outliers between robust poisson models and log-binomial models when estimating relative risks for common binary outcomes: a simulation study. BMC Medical Research Methodology 14, 1–8 (2014).

13. Barros, A. J. & Hirakata, V. N. Alternatives for logistic regression in cross-sectional studies: an empirical comparison of models that directly estimate the prevalence ratio. BMC Medical Research Methodology 3, 1–13 (2003).

14. Blizzard, L. & Hosmer, W. Parameter estimation and goodness-of-fit in log binomial regression. Biometrical Journal 48, 5–22 (2006).

15. Petersen, M. R. & Deddens, J. A. A comparison of two methods for estimating prevalence ratios. BMC Medical Research Methodology 8, 1–9 (2008).

16. Knol, M. J., Le Cessie, S., Algra, A., Vandenbroucke, J. P. & Groenwold, R. H. Overestimation of risk ratios by odds ratios in trials and cohort studies: alternatives to logistic regression. CMAJ 184, 895–899 (2012).

17. Williamson, J. M., Datta, S. & Satten, G. A. Marginal analyses of clustered data when cluster size is informative. Biometrics 59, 36–42 (2003).

18. Seaman, S., Pavlou, M. & Copas, A. Review of methods for handling confounding by cluster and informative cluster size in clustered data. Statistics in Medicine 33, 5371–5387 (2014).

19. Williamson, J. M.Kim, H.-Y. & Warner, L. Weighting condom use data to account for nonignorable cluster size. Annals of Epidemiology 17, 603–607 (2007).

20. Yland, J. et al. Methodological approaches to analyzing IVF data with multiple cycles. Human Reproduction 34, 549–557 (2019).

21. Dodge, L. E. et al. Choice of statistical model in observational studies of ART. Human Reproduction 35, 1499–1504 (2020).

22. Chaurasia, A., Liu, D. & Albert, P. S. Pattern–mixture models with incomplete informative cluster size: application to a repeated pregnancy study. Journal of the Royal Statistical Society Series C: Applied Statistics 67, 255–273 (2018).

23. Wang, M., Kong, M. & Datta, S. Inference for marginal linear models for clustered longitudinal data with potentially informative cluster sizes. Statistical Methods in Medical Research 20, 347–367 (2011).

24. Mitani, A., Kaye, E. & Nelson, K. Marginal analysis of multiple outcomes with informative cluster size. Biometrics 77, 271–282 (2021).

25. Iosif, A.-M. & Sampson, A. R. A model for repeated clustered data with informative cluster sizes. Statistics in Medicine 33, 738–759 (2014).

26. Dasa, T. T., Okunlola, M. A. & Dessie, Y. Effect of grand multiparity on the adverse birth outcome: A hospital-based prospective cohort study in Sidama Region, Ethiopia. International Journal of Women’s Health, 363–372 (2022).

27. Hoffman, E. B., Sen, P. K. & Weinberg, C. R. Within-cluster resampling. Biometrika 88, 1121–1134 (2001).

28. Wang, Z. et al. Early-life menstrual characteristics and gestational diabetes in a large US cohort. Paediatric and Perinatal Epidemiology 38, 654–665 (2024).

29. Wilson, J. E., Sanderson, W., Westgate, P. M., Winter, K. & Forster, D. Risk factors of carbapenemase-producing Enterobacterales acquisition among adult intensive care unit patients at a Kentucky Academic Medical Center. Infection Prevention in Practice 5, 100310 (2023).

30. Hansen, K. R. et al. Intrauterine insemination performance characteristics and post-processing total motile sperm count in relation to live birth for couples with unexplained infertility in a randomised, multicentre clinical trial. Human Reproduction 35, 1296–1305 (2020).

31. Craig, L. B. et al. Racial and ethnic differences in pregnancy rates following intrauterine insemination with a focus on American Indians. Journal of Racial and Ethnic Health Disparities 5, 1077–1083 (2018).

32. Edmonds, A. T. et al. Patient-centered primary care and receipt of evidence-based alcohol-related care in the national Veterans Health Administration. Journal of Substance Abuse Treatment 138, 108709 (2022).

33. Ma, Y., Wang, H. & Jiang, X. Penalized weighted GEEs for high-dimensional longitudinal data with informative cluter size. arXiv preprint 2501.00839 (2025).

34. Williamson, T., Eliasziw, M. & Fick, G. H. Log-binomial models: exploring failed convergence. Emerging Themes in Epidemiology 10, 1–10 (2013).

35. Huang, Y. & Leroux, B. Informative cluster sizes for subcluster-level covariates and weighted generalized estimating equations. Biometrics 67, 843–851 (2011).

36. Hastie, T., Tibshirani, R., Friedman, J. H. & Friedman, J. H. The elements of statistical learning: data mining, inference, and prediction (Springer, 2009).

37. Bennett, A. S. et al. Naloxone protection, social support, network characteristics, and overdose experiences among a cohort of people who use illicit opioids in New York City. Harm Reduction Journal 19, 20 (2022).

38. Elliott, L., Chen, Y., Goodman, M. & Bennett, A. S. Distal factors associated with proximal overdose risk behaviors and recent non-fatal overdose among a sample of people who use illicit opioids in New York City. Journal of Drug Issues 54, 457–475 (2024).

39. Narrow, W. E. et al. DSM-5 field trials in the United States and Canada, Part III: development and reliability testing of a cross-cutting symptom assessment for DSM-5. American Journal of Psychiatry 170, 71–82 (2013).

40. Zhang, Y. et al. Long-term participant retention and engagement patterns in an app and wearable-based multinational remote digital depression study. NPJ Digital Medicine 6, 25 (2023).

41. Lin, C., Howard, V. J., Nanavati, H. D., Judd, S. E. & Howard, G. The association of baseline depressive symptoms and stress on withdrawal in a national longitudinal cohort: the REGARDS study. Annals of Epidemiology 84, 8–15 (2023).

42. Talbot, D., Mésidor, M., Chiu, Y., Simard, M. & Sirois, C. An alternative perspective on the robust Poisson method for estimating risk or prevalence ratios. Epidemiology 34, 1–7 (2023).

43. Huang, F. L. Analyzing cross-sectionally clustered data using generalized estimating equations. Journal of Educational and Behavioral Statistics 47, 101–125 (2022).

44. Wilms, R., Mäthner, E., Winnen, L. & Lanwehr, R. Omitted variable bias: A threat to estimating causal relationships. Methods in Psychology 5, 100075 (2021).

45. Mancl, L. A. & DeRouen, T. A. A covariance estimator for GEE with improved small-sample properties. Biometrics 57, 126–134 (2001).

46. Kauermann, G. & Carroll, R. J. A note on the efficiency of sandwich covariance matrix estimation. Journal of the American Statistical Association 96, 1387–1396 (2001).

47. Fay, M. P. & Graubard, B. I. Small-sample adjustments for Wald-type tests using sandwich estimators. Biometrics 57, 1198–1206 (2001).

48. Benhin, E., Rao, J. & Scott, A. Mean estimating equation approach to analysing cluster-correlated data with nonignorable cluster sizes. Biometrika 92, 435–450 (2005).

49. Lee, D., Kim, J. K. & Skinner, C. J. Within-cluster resampling for multilevel models under informative cluster size. Biometrika 106, 965–972 (2019).

50. Nevalainen, J., Oja, H. & Datta, S. Tests for informative cluster size using a novel balanced bootstrap scheme. Statistics in Medicine 36, 2630–2640 (2017).

51. Anyaso-Samuel, S., Datta, S., Roos, E. & Nevalainen, J. Can the unit size predict outcomes? Testing for informativeness in three-level designs. Statistics in Medicine 44, e70041 (2025).

52. Kim, S., Martens, M. J. & Ahn, K. W. Test statistics and statistical inference for data with informative cluster sizes. Biometrical Journal 67, e70021 (2025).

